# Shared and Sex-Specific Genetic Risk for Parkinson’s Disease Risk Across European Populations

**DOI:** 10.64898/2025.12.29.25343160

**Authors:** The Global Parkinson’s Genetics Program (GP2), Hampton L. Leonard

## Abstract

**Introduction:** Parkinson’s disease (PD) affects females and males differently, with differences in prevalence, clinical phenotypes, and therapeutic response, suggesting that biological sex may influence the underlying molecular mechanisms of PD. The extent to which genetic factors may contribute to these differences remains largely unknown.

**Objective:** To investigate sex-specific autosomal genetic factors associated with PD risk.

**Methods:** We performed a sex-stratified autosomal GWAS meta-analysis leveraging data from the Global Parkinson’s Genetics Program, the International Parkinson’s Disease Genomics Consortium, the UK Biobank, and the Fox Insight Genetics Study, including a total of 226,196 individuals from different European populations: 18,145 female PD cases, 95,558 female controls, 28,747 male PD cases, and 83,746 male controls.

**Results:** We observed a high genetic correlation between the male and female PD meta-analyses (rg = 0.909, SE = 0.0403; p = 8.03E-113), and the heritability estimates were comparable between sexes (∼10% in males and ∼11% in females) and similar to estimates from prior sex-combined analyses. Our sex-stratified GWAS identified 57 genome-wide significant association signals, including five novel risk loci, three of which reached genome-wide significance in males only (*RBM8A, ANKRD23,* and *CNTN4*) and two in females only (*RERE* and *ARL6IP6*). Of the remaining previously identified GWAS loci regions, several showed differences in effect magnitude between sexes, with the *GALC, RERE, ARL6IP6,* and *RBM8A* loci demonstrating statistically significant sex-specific effects.

**Conclusions:** Overall, PD genetic architecture appears broadly similar between females and males, but the identification of five novel loci and significant differences at select regions highlights the value of sex-stratified analyses for uncovering additional genetic contributors to PD risk beyond those detected in combined analyses.

## INTRODUCTION

Parkinson’s disease (PD) is a multifactorial neurodegenerative disorder that affects females and males differently, with well-documented differences in prevalence, clinical progression, neuropsychiatric manifestations, and therapeutic response. These observations suggest that biological sex may influence the underlying molecular mechanisms of PD ^1–3^. Yet the origin of these disparities remains poorly understood and may be complex, including environmental and lifestyle factors, age-related influences, or potential genetic contributors. In this context, genetics has emerged as a key area of interest for understanding sex-related variation in PD, and growing evidence points to several ways in which genetics may contribute to these differences.

For example, some studies have detected pleiotropy, in which genetic loci affect both PD and female-specific traits such as age at menarche and menopause ^4^. Mendelian randomization analyses suggested that variants linked to a later age at menopause are associated with a reduced PD risk, suggesting neuroprotective effects of sex hormones ^5^.

Sex differences in PD at the level of gene expression and epigenetic regulation have also been reported. A meta-analysis exploring sex-specific and sex-dimorphic gene expression changes in people with PD identified 36 female-specific genes, 539 male-specific genes, and 37 genes with opposite expression patterns between females and males ^6^. Furthermore, genes involved in neuronal development, neurotransmitter regulation, and neuroprotection, such as *PARK7*, *NR4A2*, *SLC17A6*, and *PTPRN2,* exhibit sex-specific changes in DNA methylation ^7^.

Regarding genetic associations with PD risk, only a few sex-stratified genome-wide association studies (GWAS) have separately evaluated females and males with PD. The largest prior sex-stratified GWAS (13,020 male PD cases, 7,936 paternal proxy cases, 89,660 male controls; 7,947 female PD cases, 5,473 maternal proxy cases, and 90,662 female controls.) performed in European populations identified 19 genome-wide significant loci, but found no statistically significant sex-specific effects ^8^. Another substantially smaller GWAS performed in the Korean population reported potential sex-dependent effects at loci such as *SNCA*, *PARK16*, and *LRRK2* ^9^, suggesting that the sex-specific genetic architecture might vary across ancestries.

Overall, despite well-documented sex differences in PD prevalence and progression, existing GWAS have provided limited evidence for differences in risk allele effects between males and females, likely reflecting constraints in sample size and statistical power. This underscores the need for larger and more comprehensive sex-stratified analyses to clarify potential sex-related patterns in PD genetic risk. Our study addresses this gap by conducting the largest sex-stratified PD GWAS meta-analysis in European populations to date, leveraging large-scale genetic data from the Global Parkinson’s Genetics Program (GP2), the International Parkinson’s Disease Genomics Consortium (IPDGC), the UK Biobank (UKB), and the Fox Insight Genetics Study (FIGS) to investigate shared and sex-dependent patterns of genetic susceptibility.

## METHODS

### Study design

Our study employed a sex-stratified design to identify genetic variants associated with PD, conducting all analyses separately for males and females (**Figure 1**). We analyzed data from individuals of European ancestry (EUR) and Ashkenazi Jewish ancestry (AJ), incorporating two types of study designs: traditional case-control cohorts with clinically diagnosed cases based on the Queen Square Brain Bank Criteria or similar diagnostic criteria (GP2 and IPDGC), and population-based biobank cohorts using electronic medical records with ICD-10 codes or proxy case status (Fox Insight and UK Biobank).

**Figure 1:**
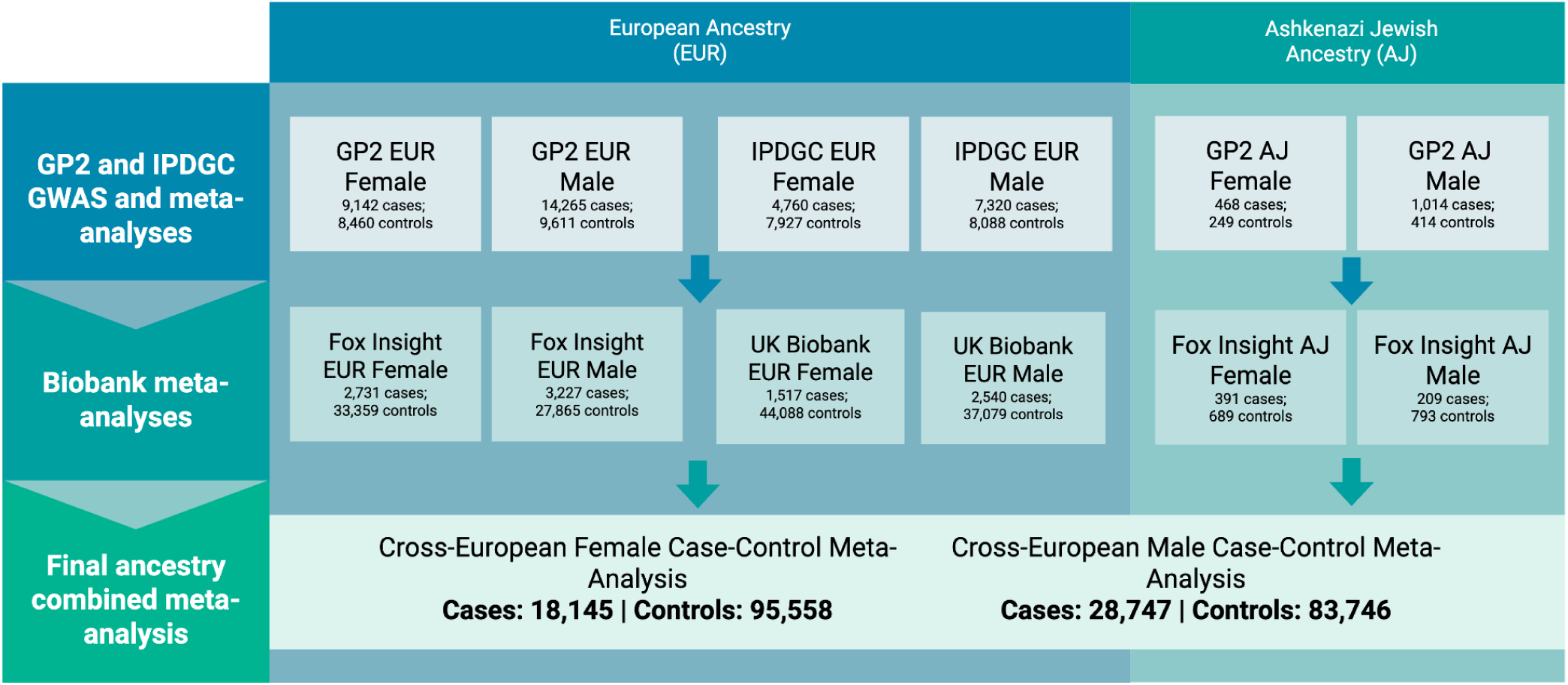
Flowchart detailing study design schema and sample counts. Study design and sample size for each ancestry and dataset. Refer to Supplementary Table 1 for a more detailed breakdown of sample size per dataset.

For each sex, we conducted separate meta-analyses within each ancestry group (EUR and AJ) and study type (case-control and biobank). These were then combined into cross-European meta-analyses, which served as the primary analyses for downstream investigations. Following meta-analysis, we applied stringent quality control filters to ensure robust associations: heterogeneity (I²) < 80, minor allele frequency (MAF) > 0.01 (with exceptions for known PD-associated genes: *GBA1*, *LRRK2*, and *SNCA*), and presence in >50% of contributing studies (≥8 studies).

Variants passing these filters were prioritized for follow-up analyses, including: (1) quality control procedures such as T-tests to identify potentially sex-specific loci and control-control GWAS to exclude artifacts unrelated to PD; (2) functional annotation and gene-based analyses using FUMA and MAGMA to assess gene enrichment in tissues and cell types; and (3) functional inference through colocalization analysis with brain eQTLs and linkage disequilibrium score regression (LDSC) to estimate heritability and genetic correlation.

### Datasets analyzed

For a complete inventory of all datasets analyzed, please refer to **Supplementary Table 1**. The majority of cases were sourced from GP2, genotyped on the NeuroBooster Array ^10^. Additional non-overlapping samples were obtained from datasets curated by the IPDGC ^11,12^, comprising 10 independent cohorts genotyped on various platforms, including NeuroChip, Illumina HumanHap550, Illumina Human OmniExpress, and Expanded Multi-Ethnic Genotyping Array. These studies comprised the traditional clinical case-control study portion of this analysis, as their diagnoses were primarily based on the Queen Square Brain Bank Criteria as part of recruitment. Biobank-scale data from the UKB were genotyped on the UKB Axiom Array and UK BiLEVE Axiom Array. A case-control GWAS defined by ICD coding criteria was conducted using UKB data. Cases from the Fox Insight Genetics Study (FIGS), which includes a subset of the 23andMe PD samples, were matched to aged population controls with comparable Illumina arrays and processed in the same manner as the GP2 samples using GenoTools^13–15^. The UKB and FIGS cohorts comprised the biobank portion of this analysis.

### Cohort quality control and ancestry

GP2, FIGS, and each non-overlapping IPDGC dataset underwent standard quality control and ancestry estimation via GenoTools, described in detail elsewhere ^16^. As there is individual overlap between the IPDGC studies and GP2, in addition to the standard relatedness filtering within studies performed by GenoTools, duplication and relatedness to GP2 samples were assessed using KING ^17^ for each IPDGC dataset. Samples that were duplicated or related to GP2 samples on a level closer than first-degree cousins were removed from the IPDGC studies, to prioritize GP2 samples, which were all genotyped on the NeuroBooster Array ^10^. Only predicted ‘EUR’ samples were used from the IPDGC data, as predicted ‘AJ’ ancestry sample sizes were too small (n<100 cases or controls). To construct the combined FIGS and population control dataset, we kept FIGS cases from the largest sample set that had been genotyped on the same Illumina Global Screening Array and merged these with population controls genotyped on the same array from the National Cancer Institute (NCI). NCI samples were labeled as population controls if they were both free from any neurological disease and were not taking a PD drug. After QC with GenoTools, each dataset was imputed separately by genetically predicted ancestry using the TOPMed-r3 reference panel. Imputed data was then filtered for imputation quality greater than 0.3 prior to analyses.

UKB quality control was performed previously ^18^, however, to keep ancestry prediction methods similar, the GenoTools ancestry prediction module was run on the UKB genotyped samples. Samples predicted ‘EUR’ were then used for the case control and the proxy analyses (sample sizes for predicted ‘FIN’ and ‘AJ’ ancestries were too small). PD cases were defined by the UKB algorithmically-defined code p42032, which includes ICD9, ICD10, and self-report codes curated by UKB. Controls were sampled at 20X the case sample size from anyone currently older than 60 years of age who did not have a value for p42032, did not have any reported ICD10 codes of G20, G21, G22, G23, G24, G25, and did not report family history of PD. Proxy cases were defined as anyone who reported a mother or father with PD. Proxy controls were selected using the same criteria as used for the case-control analysis. Related individuals were removed prior to analysis.

### GWAS and meta-analyses

GWAS were conducted using Plink2 ^19^, correcting for principal components, genetically-determined sex, and age (when available). Summary statistics were filtered for minor allele frequency (MAF) > 1% specific to each study, except for the *GBA1*, *SNCA*, and *LRRK2* regions (gene ±250kb). Fixed-effect and random-effects meta-analyses were carried out per ancestry and per case ascertainment criteria (clinical case-control or biobank), as described, and then, finally, for the full joint analysis using Plink1.9 ^20^. For each meta-analysis, we defined loci only using variants that were filtered for quality by keeping variants that were present in at least 50% of our total studies (8 out of 15) and had an I^2^ heterogeneity estimate < 80. Genomic inflation estimates were calculated for each meta-analysis. Scaled genomic inflation was nominal and at or below 1.03 for all meta-analyses (with the exception of the AJ female meta-analysis, which lacked the sample size for effective lambda_1000_ scaling), all lambda and scaled lambda_1000_ estimates ^21^ can be found in **Supplementary Table 2**. Significant independent loci were determined by the genome-wide significance threshold of 5e-8 and by using a linkage disequilibrium (LD) clumping threshold of r^2^ > 0.6 within 250kb windows, facilitated by Functional Mapping and Annotation (FUMA) as described elsewhere previously ^22^. A locus was classified as novel if it fell outside of a region (defined by the index variant ±250 kb) previously identified by Nalls et al., Foo et al., or Kim et al.^11,22,23^. All independent variants, known and novel, can be found in **Supplementary Table 3**.

The discovery results for the meta-analyses were run using GP2 data from release 9. As a replication cohort, we used the non-overlapping European-ancestry samples added to GP2 in release 10, ensuring that only independent participants not included in the discovery analysis were analyzed to investigate the lead-nominated variants for each sex. Association tests were performed in the same way as for the discovery meta-analyses. The replication analysis consisted of 1,634 cases and 1,026 controls for females and 3,026 cases and 1,057 controls for males.

To assess whether observed differences reflected statistically significant sex-specific effects, we performed two-sample Z-tests comparing effect size estimates between males and females. Analyses were conducted separately for males and females and restricted to variants that were significant in one sex while the corresponding lead variant in the region in the other sex had a P-value > 5e-6. Resulting P-values were Bonferroni-corrected to account for multiple testing.

To ensure our results did not contain sex-specific artifacts, we ran a male versus female GWAS in controls only in the GP2 cohort. The control GWAS was also conducted with Plink2 and corrected for principal components and age (when available). This analysis was run in EUR ancestry only for sample-size purposes, and consisted of 9,611 male controls and 9,142 female controls.

### Genetic correlation and heritability

Genetic correlations and heritability were calculated using summary statistics with linkage disequilibrium score regression (LDSC) under default settings. We investigated the correlation between sex-specific meta-analyses and between biobank and case-control sex-specific meta-analyses, as well as between sex-specific AJ and EUR ancestry meta-analyses. LDSC was run 2 times, one assuming the same disease prevalence across cases of roughly 2% as cited in previous literature, and another assuming different prevalence estimates per sex (0.6% for females and 1% for males) as previously reported ^24^.

### Colocalization analysis and prioritization

We performed Bayesian colocalization analysis using the COLOC method to identify genes whose expression may mediate genetic associations with PD risk. Sex-stratified GWAS summary statistics were used alongside expression quantitative trait loci (eQTL) data from GTEx v10 across 13 brain tissues: amygdala, anterior cingulate cortex (BA24), caudate nucleus (basal ganglia), cerebellar hemisphere, cerebellum, cortex, frontal cortex (BA9), hippocampus, hypothalamus, nucleus accumbens (basal ganglia), putamen (basal ganglia), spinal cord (cervical c-1), and substantia nigra. For each gene-tissue-sex combination, COLOC computed posterior probabilities for five hypotheses: H0 (no association with either trait), H1 (association with GWAS only), H2 (association with eQTL only), H3 (association with both traits via distinct causal variants), and H4 (shared causal variant between traits).

To prioritize high-confidence colocalization signals, we applied a multi-tiered filtering approach based on established best practices. Gene-tissue pairs were classified as "Strong" evidence if PP.H4 ≥ 0.80, "Suggestive" evidence if 0.50 ≤ PP.H4 < 0.80, or "None" otherwise. Additional quality control filters were implemented to reduce false positives: (1) signal evidence filters requiring (PP.H1 + PP.H4) ≥ 0.80 and (PP.H2 + PP.H4) ≥ 0.80 to ensure both GWAS and eQTL signals were present in the region; (2) PP.H3 ≤ 0.20 to exclude loci with evidence for distinct causal variants; (3) PP.H0 ≤ 0.10 to exclude loci lacking association evidence; and (4) a minimum of 50 SNPs tested in the colocalization region to ensure adequate statistical power. Gene-tissue pairs failing any quality control criterion were excluded from downstream analysis regardless of their PP.H4 value.

We further stratified results by sex-specific and sex-shared signals, enabling identification of genes with colocalization evidence in both sexes, male-only, or female-only contexts.

## RESULTS

### Summary of findings

The final dataset for the meta-analysis, combining data from all studies and different European populations, comprised 226,196 individuals, including 18,145 female PD cases, 95,558 female controls, 28,747 male PD cases, and 83,746 male controls (**Supplementary Table 1**). The sex-stratified GWAS identified 57 genome-wide significant association signals, displayed in the Miami plot in **Figure 2**. We identified five previously unreported risk loci, three of which reached genome-wide significance in males only (*RBM8A, ANKRD23,* and *CNTN4*) and two in females only (*RERE* and *ARL6IP6*). Of the remaining 52 known loci, 25 were genome-wide significant in both sexes, while the others reached genome-wide significance in females or males only (**Supplementary Table 3**). Among the 25 shared loci, five (*GBA1, TMEM175, SNCA, BAG3,* and *LRRK2*) showed the identical lead variants in males and females, while the remaining loci were represented by distinct lead SNPs across sexes.

**Figure 2:**
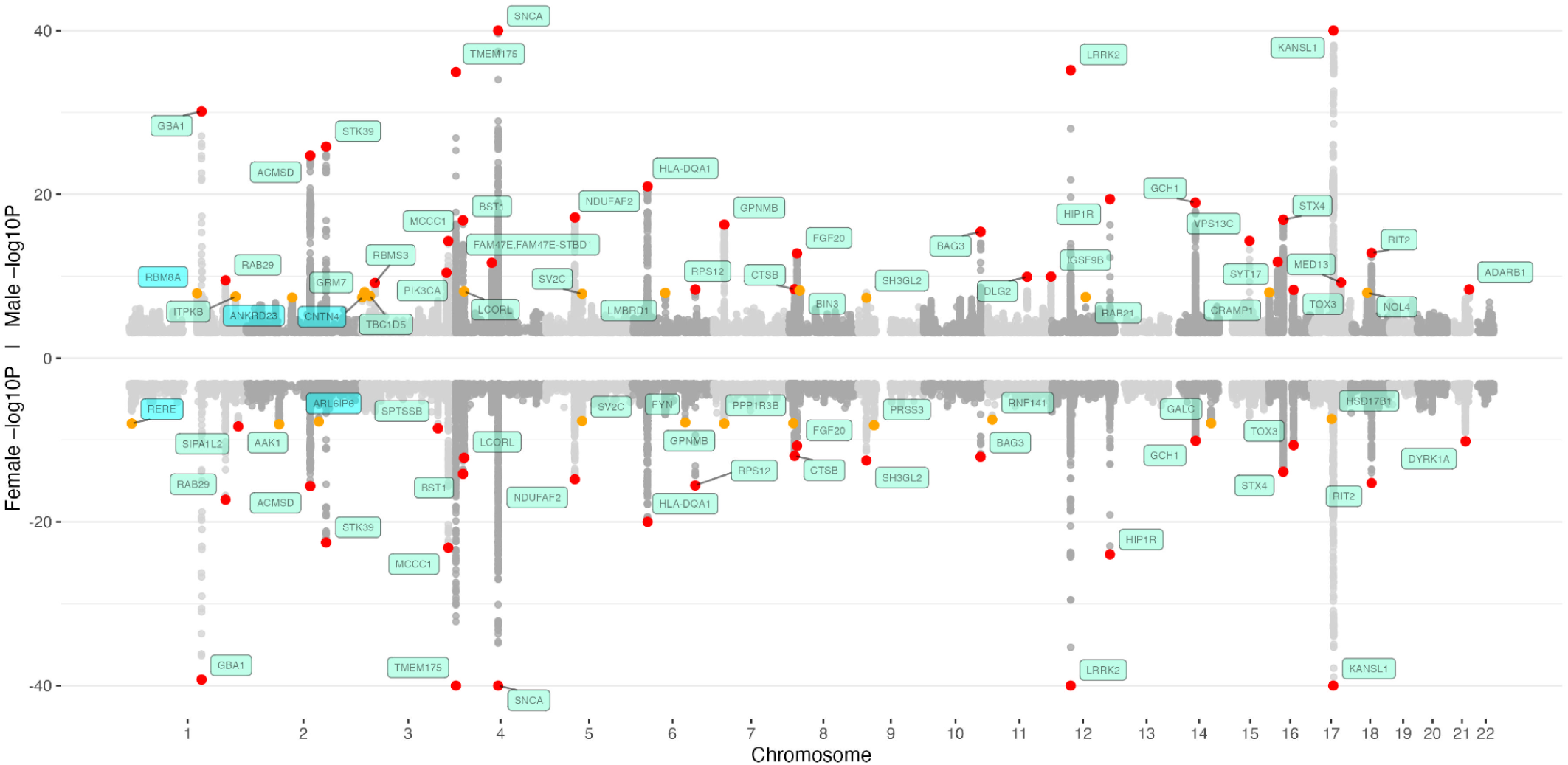
Miami plot for male and female meta-analyses. The nearest gene to each of the 57 significant loci is labeled in green for previously-identified loci and in blue for novel loci. −log10 P values were capped at 40. Variant points are color-coded, orange indicates significance levels between P < 5E-8 and P = 5E-9, while red indicates variants with P < 5E-9.

### Identification of novel and independent loci in males and females across European populations

Five novel loci were identified in our sex-stratified European cross-study meta-analysis, meaning that these loci were not previously reported in any PD GWAS. Three of which reached genome-wide significance in males only and two in females only, underscoring the added value of sex-specific analyses for uncovering novel genetic contributors to PD. In males, genome-wide significant associations were detected near *RBM8A*, driven by rs71582802 (chr1:145928008:T:A; beta = -0.1241, SE = 0.021794, P = 1.24E-8), near *ANKRD23* with rs11897786 (chr2:96841135:G:A; beta = -0.1204, SE = 0.021942, P = 4.08E-8), and near *CNTN4*, where rs9827746 (chr3:2182935:C:G) reached genome-wide significance (beta = -0.0655, SE = 0.011911, P = 3.82E-8). None of these loci showed a corresponding genome-wide significant signal in females. However, some level of association was observed in females at these five loci, but not for the same underlying signal. For example, in the *RBM8A* region, the lead SNP in males (rs71582802) did not show a P-value of interest in females (P = 5.50E-01). However a different SNP (rs115243157, R2 = 0.0031, D’ = 1) was nominated as the lead SNP in females in the same region where some significance was identified (P = 7.27E-03).

Conversely, genome-wide significant associations in females but not males were observed at two independent loci: near *RERE* with rs301802 (chr1:8437247:T:A; beta = -0.0785, SE = 0.01371, P = 1.03E-8) and near *ARL6IP6* with rs7607599 (chr2:152862205:C:G; beta = -0.0782, SE = 0.013879, P = 1.76E-8). Similarly, independent SNPs in the same region for males show some level of significance. Full effect estimates for all novel genome-wide significant SNPs across both sexes are summarized in **Table 1**.

Additionally, there was one novel signal observed in the AJ ancestry female stratified meta-analysis. rs4916793 (chr5:90261068:C:T) is located in an intron of long non-coding RNA LINC01339. However, the random effects p-value for this variant did not pass the genome-wide significance threshold, suggesting study heterogeneity (P = 0.0001622). This locus will need to be replicated before further conclusions can be drawn. All genome-wide significant loci nominated in the AJ population can be found in **Table 2**.

In the male versus female control GWAS, no loci were significant at 5E-8, including the novel loci highlighted in **Table 1** and the differential effect loci in **Table 3**. This suggests that our results nominated in the sex-stratifed PD meta-analyses are not driven by underlying biological differences associated with sex but are potentially driven by disease differences. 11 loci passed a reduced threshold of 5e-6, but none of these 11 loci were within ±250kb of the loci nominated in the stratified analyses. In addition, we also compared our results with the results from Pirastu et al., which performed a large GWAS of sex in over 2 million participants from 23andMe^25^. None of our novel loci in **Table 1** or the differential effect loci in **Table 3** were within ±250kb of the 158 variants nominated in Pirastu et al. This provides further evidence that our results are not due to underlying biological differences or sex-related participation bias in cohorts.

### Variation in known PD risk loci between males and females

While several of the previously identified PD risk loci reached genome-wide significance in both males and females, others showed notable differences between sexes, either related to statistical significance, direction of effect, or the lead variant identified. To test whether some of these differences translated to significant sex-specific effects, we selected variants significant in one sex where the lead variant in the same locus in the other sex had a P-value > 5E-6. This resulted in 12 loci for males and 10 loci in females. In males, 4 loci had a significantly different magnitude of effect from females (*RBM8A*, *ANKRD23*, *CNTN4*, *BIN3*) but only *RBM8A* passed the male multiple test-correction threshold for 12 tests. In females, 9 loci had a significantly different magnitude of effect from females (*RERE*, *SIPA1L2*, *AAK1*, *ARL6IP6, FYN, PPP1R3B, PRSS3, RNF141, GALC*) and 3 loci passed multiple test correction including *RERE*, *ARL6IP6*, and *GALC*.

At the *GALC* locus the lead SNP in females, rs3213916 (chr14:87949673:G:A), reached genome-wide significance exclusively in females (beta = 0.081, SE =0.014178, P = 1.11E-8), whereas no significant association was observed in males (beta = 0.0224, SE = 0.0124, P = 0.071). Further, in the male-only analysis, rs59884956 (chr14:87905950:G:C, beta = -0.0407, SE = 0.012066, P = 7.44E-4) emerged as the lead SNP but was not significant and had an opposite direction of effect than the female lead SNP. Interestingly, similar, albeit smaller, differences in effect estimates at the *GALC* locus have been observed before by Blauwendraat and colleagues, although their identified signal at *GALC* (rs979812) did not reach genome-wide significance in either sex (males: beta = 0.031, SE = 0.0168, P = 0.06462; females: beta = 0.0895, SE = 0.0192, P = 3.07E-6) and the observed variation between sexes was not significant after correction for multiple testing. Importantly, when comparing the beta estimates for the index variant of the most recent PD risk GWAS, it becomes clear that the female risk beta (0.071) is more than double compared to the male risk beta (0.031) and the males appear to dilute the combined risk beta from the most recent EUR risk GWAS GP2 et al (0.044)^26^. A comparison of local architecture and a locus compare plot between males and females at the *GALC* locus can be found in **Figure 4** and **Supplementary Figure 1**.

**Figure 3:**
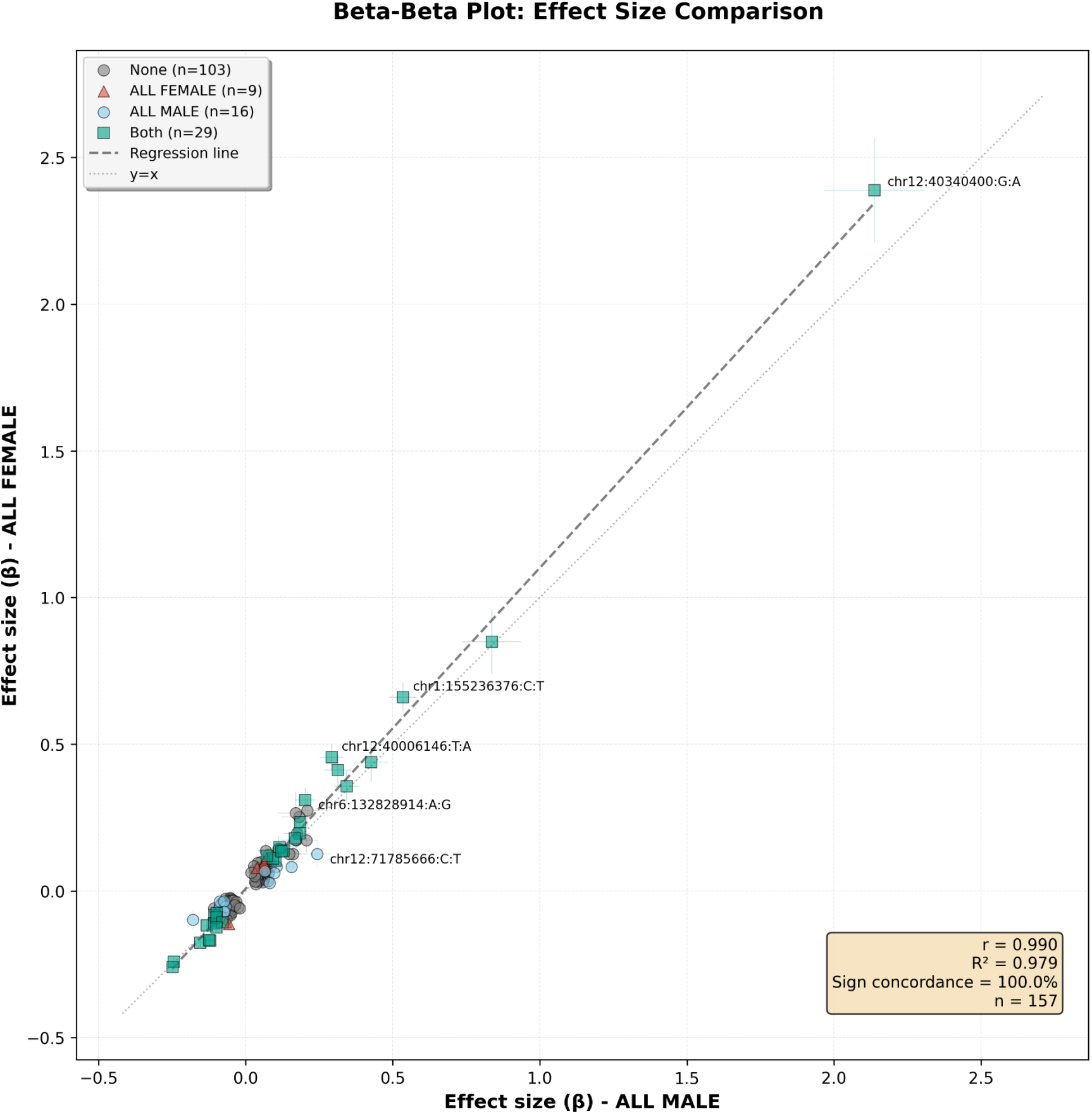
Beta-beta plot comparing effects from the male meta-analysis and the female meta-analysis using the 157 variants nominated by the most recent GP2 PD risk GWAS (2025) PD case-control meta-analysis.

**Figure 4:**
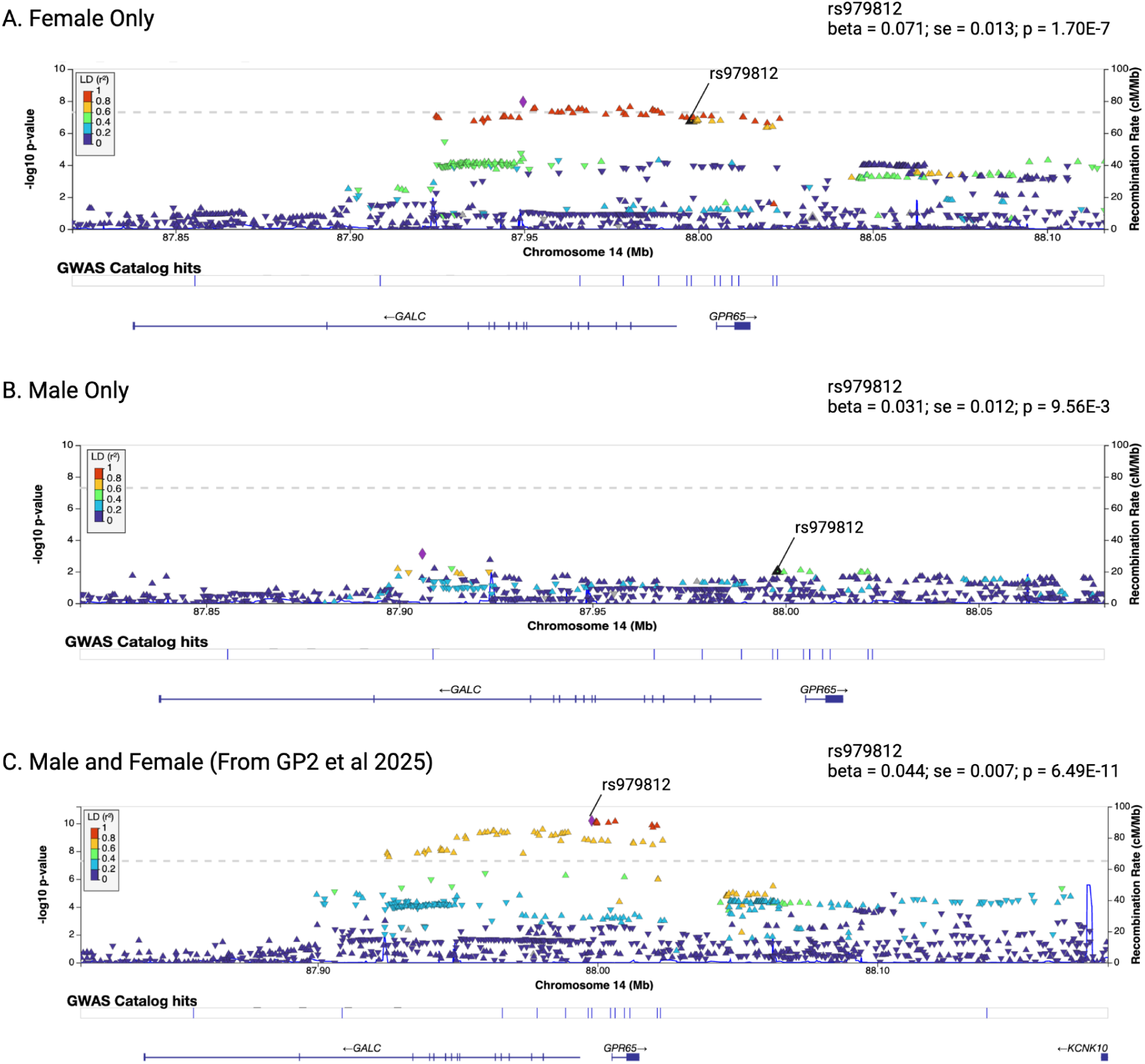
LocusZoom plots of the *GALC* locus in the female-only meta-analysis, the male-only meta-analysis, and combined male and female analysis from GP2 et al 2025.

Interestingly, the additional loci that pass test correction for significant difference in effect magnitude (*RBM8A, RERE, ARL6IP6*) are novel loci nominated in this study. These loci did not reach genome-wide significance in the most recent EUR combined risk meta-analysis, but did trend towards significance (*RBM8A:* beta = -0.0418, SE = 0.0129, P = 1.22E-03, *RERE:* beta = -0.0264, SE = 0.0068, P = 1.03E-04, *ARL6IP6:* beta = -0.0195, SE = 0.007, P = 5.03E-03). The difference in magnitude of effect for these loci follow a similar pattern as the *GALC* locus, with some even demonstrating an opposite direction of effect between sexes, suggesting the possibility that these differences may have diluted the ability to detect these loci in the combined risk meta-analysis (*RBM8A:* EUR risk GWAS beta = -0.0418, Male beta = -0.1241, Female beta = -0.0149, *RERE:* GP2 et al beta = -0.0264, Male beta = 0.0078, Female beta = -0.0785, *ARL6IP6:* GP2 et al beta = -0.0195, Male beta = 0.0038, Female beta = -0.0782). Locus compare plots between males and females for these loci can be found in **Supplementary Figures 2**, **3**, and **4**.

Detailed effect estimates for all genome-wide significant loci for both sexes are provided in **Supplementary Table 3**.

### Genetic correlation and heritability are similar between sexes

The genome-wide genetic correlation from LDSC between the combined EUR ancestry male and female PD meta-analyses was high (rg = 0.909, SE = 0.0403; P = 8.03E-113), similar to previous estimates from Blauwendraat et al (rg = 0.877, SE = 0.0699; P = 4.24E-36), indicating again that the autosomal genetic architecture of PD is largely shared between men and women. Correlation was comparable between sexes across different PD case definitions, including case-control cohorts in IPDGC and GP2 (rg = 0.8704) and Biobank cohorts (rg = 0.9529). Sex-specific prevalence estimates did not significantly affect the genetic correlation results compared to population-level prevalence estimates.

Liability-scaled heritability estimates were also similar across sexes (h2_male = 0.0994, SE = 0.0082; h2_female = 0.1107, SE = 0.011), similar to what was reported in sex-combined estimates in GP2 et al (h2 = 0.0862, SE = 0.0062). Heritability estimates were also similar between sexes when comparing both case-control cohort meta-analyses (h2_male = 0.1575, SE = 0.0144; h2_female = 0.1609, SE = 0.0185) and biobank cohort meta-analyses (h2_male = 0.1232, SE = 0.0172; h2_female = 0.1379, SE = 0.0207).

We also investigated the correlation of the signals between the male and female meta-analyses using the 157 risk signals from the GP2 et al 2025 PD case-control meta-analysis (**Figure 3**). Very high correlation was observed between the effect sizes of the sex-specific GWAS and the 157 variants (R2 > 0.99), although lead variants between sexes often differed and some, such as the above mentioned *GALC* locus, showed difference in the magnitude of effect (rs3213916, male_beta = 0.022, female_beta = 0.081).

### Colocalization of loci with eQTL data reveals shared and sex-specific effects

To identify loci with potential sex-specific expression effects, we performed colocalization fine-mapping between our GWAS signals and brain eQTLs, focusing on loci with strong evidence (PP.H4 ≥ 0.80) in one or both sexes.

Five loci that reached genome-wide significance only in females also showed female-specific colocalization with brain eQTLs. Fine-mapping identified compelling candidate genes at each locus: *RERE/RERE-AS1* (chr1:8314193:T:A, PP.H4 = 0.868 - 0.990), *AAK1/GFPT1* (chr2:69303968:G:C, PP.H4 = 0.869 - 0.949), PRSS3 (chr9:34065575:C:T, PP.H4 = 0.965), *AMPD3* (chr11:10654191:A:G, PP.H4 = 0.985), and *GALC* (chr14:87905950:G:C, PP.H4 = 0.918). These colocalization signals were observed across multiple brain regions including cerebellum, hippocampus, and frontal cortex.

Similarly, four loci significant only in males demonstrated male-specific colocalization patterns. Fine-mapping prioritized *NUDT17* (chr1:145928008:T:A, PP.H4 = 0.967), *ADORA2B* (chr2:96841135:G:A, PP.H4 = 0.894), *EME2* (chr16:1666009:A:C, PP.H4 = 0.983), and *BRIP1* (chr17:62109174:G:A, PP.H4 = 0.917) as likely causal genes at their respective loci, with signals primarily in basal ganglia and cerebellum regions.

While some loci like *RAB29* showed robust colocalization in both sexes (PP.H4 > 0.99 in both), fine-mapping also revealed more subtle sex-specific patterns. For instance, within the same locus, *BIN3* and *GPNMB* showed particularly strong male-specific colocalization (PP.H4 = 0.968 - 0.976).

These fine-mapping results suggest that some sex-specific genetic associations may potentially be driven by corresponding sex-specific gene expression differences in relevant brain tissues. All colocalization results can be found in **Supplementary Table 7.**

### Replication of identified loci in additional data

For additional replication we used non-overlapping samples in GP2 release 10 data. As expected, most loci did not replicate at genome-wide significance (p<5e-8) due to the sample size differences. However, out of the 37 loci nominated at genome-wide significance in the female stratified discovery meta-analysis, 19 loci had p values < 0.05, including established PD risk loci such as *GBA1*, *SNCA*, and *TMEM175*. For males, 22 of the 45 nominated loci had p values < 0.05 in the replication dataset, although none reached genome-wide significance.

Notably, the *HIP1R* locus replicated in the female replication GWAS at genome-wide significance (p=4.21e-8), but not in the male replication GWAS. The replication p values for all nominated loci can be found in **Supplementary Table 4**.

## DISCUSSION

We here performed the largest sex-stratified PD GWAS meta-analysis across European populations. Compared to prior efforts ^8^, this dataset substantially exceeds the number of included PD cases, resulting in markedly improved power to detect potential sex-related differences in genetic risk. Our sex-stratified meta-analysis resulted in 57 genome-wide significant signals, five of which have not been associated with PD risk before and were identified in males or females only, and four that showed statistically different magnitudes of effect between sexes.

### High genetic correlation and shared heritability between sexes

Overall, as expected, we observed a highly significant genetic correlation between the male and female PD meta-analyses, similar to previous estimates from Blauwendraat and colleagues ^8^, underscoring that the autosomal genetic architecture of PD is largely shared between men and women. The liability-scaled heritability estimates were also comparable between sexes with ∼10% in males and ∼11% in females, similar to prior estimates from sex-combined analyses performed with a similar dataset (∼9%) ^26^.

Among the known loci, a substantial proportion showed consistent and comparable associations in both males and females, consistent with the observed broadly shared genetic architecture. Notably, among those “shared” loci, five showed identical lead variants in males and females, e.g., *GBA1* rs2230288*, TMEM175* rs34311866*, SNCA* rs356182*, BAG3* rs144814361, and *LRRK2* rs34637584. These loci showed robust and consistent associations across sexes, with similar effect estimates, indicating little evidence of meaningful sex-related variation at these regions, which is not surprising, given their long and well-established role in PD susceptibility. Beyond these five loci, several other regions reached genome-wide significance in both sexes but were represented by different lead variants in males and females. However, these differences likely reflect variation in statistical power rather than biological differences.

Interestingly, several loci reached genome-wide significance in only one sex. While these observations may initially suggest potential sex-specific genetic effects, most of these loci still showed similar effect estimates in the other sex, indicating these findings may be more likely driven by statistical power or slight variation in signal strength rather than true sex-related variation.

### Five novel PD risk loci discovered through sex-stratified analyses

Finally, with our increased sample size, we identified five novel loci significantly associated with PD risk. Three of which reached genome-wide significance in males only, *RBM8A* (rs71582802), *ANKRD23* (rs11897786), and *CNTN4* (rs9827746), and two in females only, *RERE* (rs301802) and *ARL6IP6* (rs7607599), emphasizing the added value of sex-specific analyses for uncovering novel genetic contributors to PD. Interestingly, two of these genes have been linked to neurological phenotypes. *CNTN4,* encoding the neuronal adhesion molecule contactin-4, which has important functions for neuronal interactions and supporting neuronal developmental processes and is expressed across multiple brain regions, has been associated with autism spectrum disorder and other psychiatric conditions ^27,28^. *RERE* has been linked to neurodevelopmental disorder with or without anomalies of the brain, eye, or heart (OMIM 616975), an autosomal dominant syndrome characterized by infancy-onset developmental delay, intellectual disability, and behavioral disorders, such as autism spectrum disorders, with sometimes additional abnormalities involving the eye, heart, and genitourinary system ^29,30^. It encodes a nuclear receptor coregulator that has functions in protein complexes modulate the transcription of target genes ^29^. While promising, these novel loci require replication and functional follow-up to determine whether they reflect true sex-related biological differences.

### *GALC* and novel loci as candidates for sex-related variation in PD risk

Although this overall pattern suggests limited evidence for strong sex-specific effects, a few loci showed pronounced differences between males and females. For example, the *GALC* locus (rs3213916) reached genome-wide significance exclusively in females, whereas the signal observed in males showed no genome-wide significant association and had opposite effect estimates. Interestingly, the *GALC* locus was already highlighted in the prior sex-specific GWAS by Blauwendraat and colleagues, where it also showed slight differences in magnitude of effect with a much stronger, although not genome-wide significant, association in females ^8^. *GALC* has been of particular interest, because similar to *GBA1*, it encodes a lysosomal enzyme, which is crucial for the catabolism of galactosphingolipids. It is well known for its association with Krabbe disease, an autosomal-recessive disorder where biallelic mutations lead to a severe decrease in GALC enzymatic activity, resulting in central and peripheral nervous system alterations, including demyelination and neurodegenerations ^31^. In addition to genetic evidence from GWAS linking *GALC* to increased PD risk ^11,26^, prior studies also showed a link between *GALC* deficiency and alpha synuclein aggregation ^31,32^, a pathological hallmark of PD. This locus requires further genetic and functional work to clarify the biological basis of this sex-specific pattern.

### Limitations

This study has several limitations. First, due to the study design and the large-scale use of imputed genotyping data, analyses were limited to variants well represented in current imputation panels, particularly restricting our ability to assess rare variation or structural changes that may contribute to sex-related differences in PD risk. Second, X and Y chromosome variants were not included, preventing evaluation of potential sex-linked genetic effects. While these sex chromosomes are clearly of interest, we consider that a separate analysis which requires different data processing and quality control pipelines, and therefore were not included here. Third, given the increased male prevalence in PD there are 10,602 more male cases included than female cases in our study, which increases the power of detection in males. Fourth, we focused here on genetic and expression data, and did not investigate other underlying biological factors or environmental interactions that may modify differences in disease risk between sex. Finally, our analyses were restricted to individuals of European ancestries, which may limit generalizability to other populations and underscores the need for sex-stratified studies in more diverse cohorts. Importantly, GP2 is working on increasing diversity in PD genetics, exemplified by their most recent data release which included over 30,000 genotyped individuals of non-European ancestry, which will enable similar studies to be conducted in these populations.

### Conclusions

Taken together, our findings indicate that the PD genetic architecture is largely similar in females and males, however, we do provide evidence for significant sex-dependent genetic effects at select loci. The identification of clear examples such as *GALC,* five novel loci, and additional subtle yet potentially meaningful differences at select loci through sex-stratified analyses highlights the added value of this approach and demonstrates that examining sexes separately can uncover additional genetic contributors to PD risk and highlight regions where patterns may differ slightly between sexes. These findings support the continued use of sex-aware analytic frameworks to refine our understanding of PD susceptibility.

## ACKNOWLEDGEMENTS

This project was supported by the Global Parkinson’s Genetics Program (GP2; https://gp2.org). GP2 is funded by the Aligning Science Across Parkinson’s (ASAP) initiative and implemented by The Michael J. Fox Foundation for Parkinson’s Research (MJFF). For a complete list of GP2 members see doi.org/10.5281/zenodo.7904831.

This research was supported by the Aligning Science Across Parkinson’s Initiative, the Intramural Research Program, National Institute on Aging, National Institutes of Health, Department of Health and Human Services, project ZO1 AG000949, and the Michael J. Fox Foundation for Parkinson’s Research. This work utilized the computational resources of the NIH STRIDES Initiative (https://cloud.nih.gov) through the Other Transaction agreement - Azure: OT2OD032100, Google Cloud Platform: OT2OD027060, Amazon Web Services: OT2OD027852. This work utilized the computational resources of the NIH HPC Biowulf cluster (https://hpc.nih.gov).

This research has been conducted using the UK Biobank Resource under Application Number 33601.

## DATA AND CODE AVAILABILITY

Data used in the preparation of this article were obtained from GP2. Specifically, we used Tier 2 data from GP2 releases 9 (https://doi.org/10.5281/zenodo.14510099) and 10 (https://doi.org/10.5281/zenodo.15748014). Tier 1 data can be accessed by completing a form on the Accelerating Medicines Partnership in Parkinson’s Disease (AMP®-PD) website (https://amp-pd.org/register-for-amp-pd). Tier 2 data access requires approval and a Data Use Agreement signed by your institution. UK Biobank data can be accessed at (https://www.ukbiobank.ac.uk/use-our-data/apply-for-access/). Fox Insight data can be accessed at (https://foxinsight.michaeljfox.org/register).

All code generated for this article, and the identifiers for all software programs and packages used, are available on GitHub (https://github.com/GP2code/GP2-Male-Female-GWAS) and were given a persistent identifier via Zenodo (DOI 10.5281/zenodo.18089825).

Summary statistics from every ancestry-level meta-analysis will be available on NDPK https://ndkp.hugeamp.org/.

## CONTRIBUTIONS

Hampton L. Leonard, Mary B. Makarious, Lara M. Lange, Paula Reyes, Kristin Levine, Dan Vitale, Nicole Kuznetsov and Mike A. Nalls designed the analysis, analyzed the data, and drafted the initial manuscript.

Kristin Levine, Hampton L. Leonard, Mary B. Makarious, Dan Vitale, Mat Koretsky, and Nicole Kuznetsov contributed to quality control of GP2 genetic data and making it available to researchers.

Cornelis Blauwendraat and Andrew B. Singleton led study design, data logistics, and funding of the study.

Huw Morris, Manuela Tan, Hirotaka Iwaki, Simona Jasaityte, Ellie Stafford, Lietsel Jones, Shannon Ballard, J Solle, and Claire Wegel (Complex and Compliance Working Groups) designed the GP2 complex study, clinical and genetic data generation and preparation, and legal / data sharing logistics.

All other members of GP2 (contributors) contributed data and made critical revisions to this article. Please refer to the GP2 Banner Authorship List for a full list of contributing members and declarations (https://doi.org/10.5281/zenodo.17753486)

## Figures and Legends

**Supplementary Figure 1:**
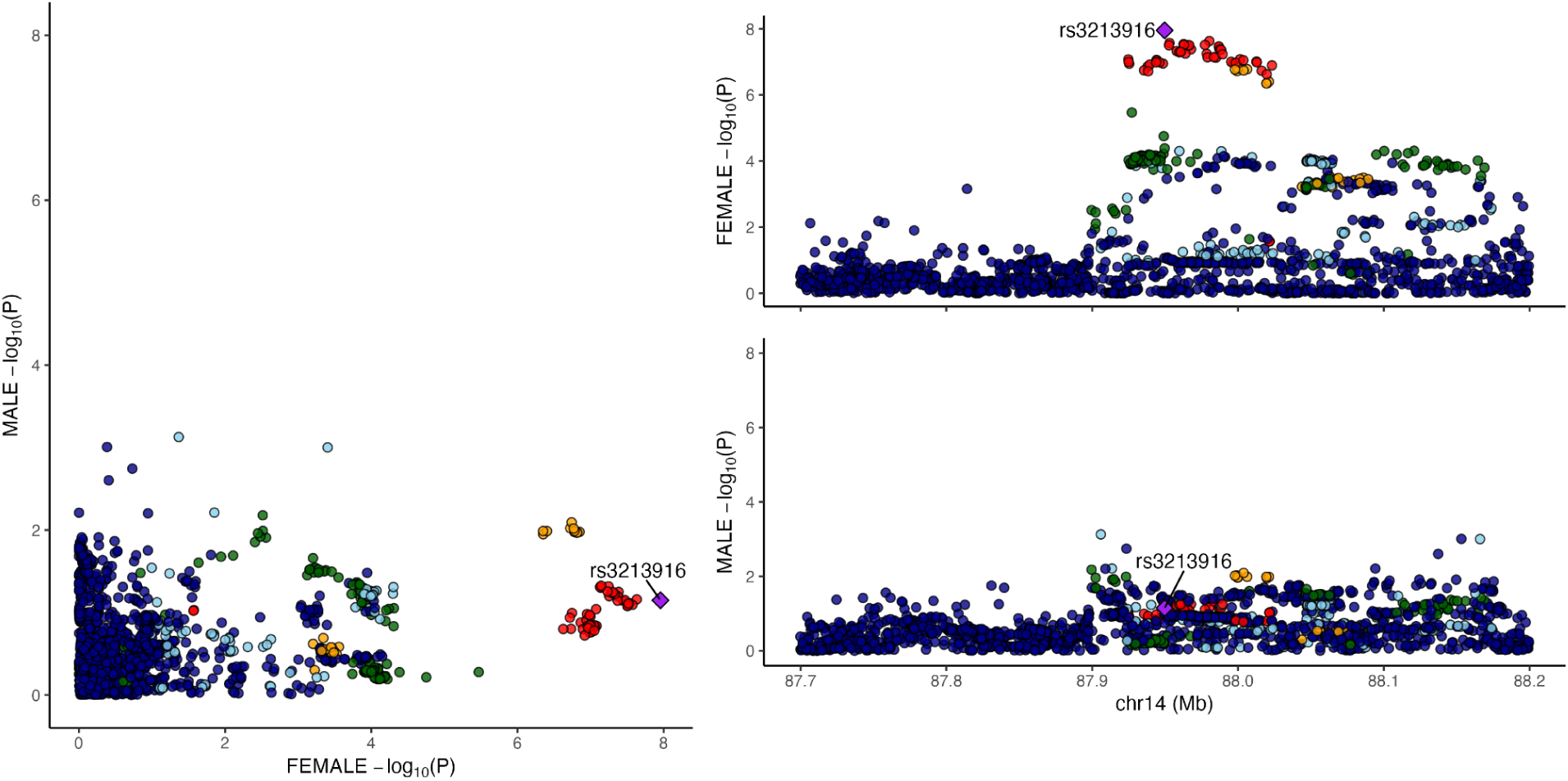
LocusCompare plot of the *GALC* locus (rs3213916).

**Supplementary Figure 2:**
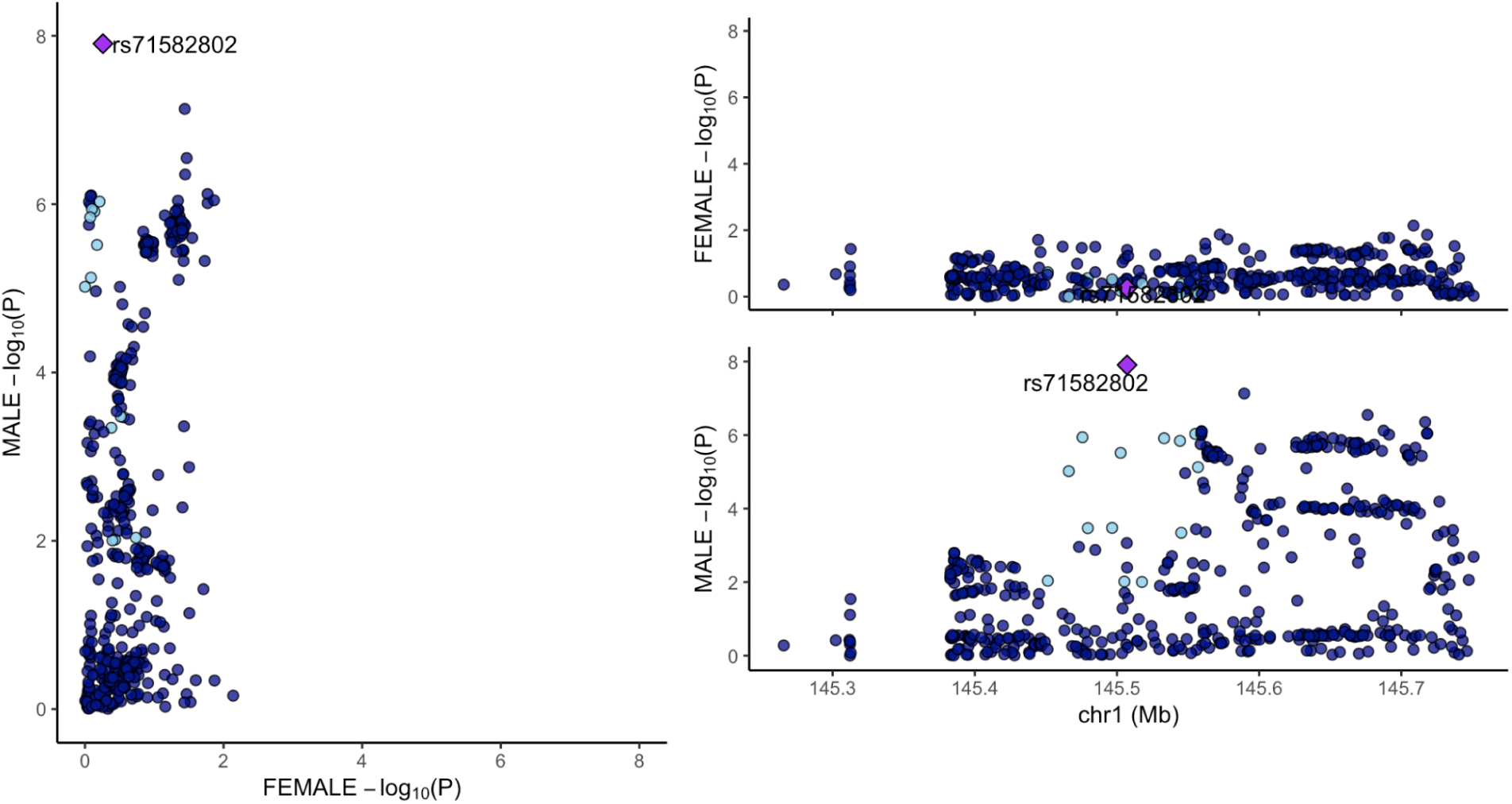
LocusCompare plot of the *RBM8A* locus (rs71582802).

**Supplementary Figure 3:**
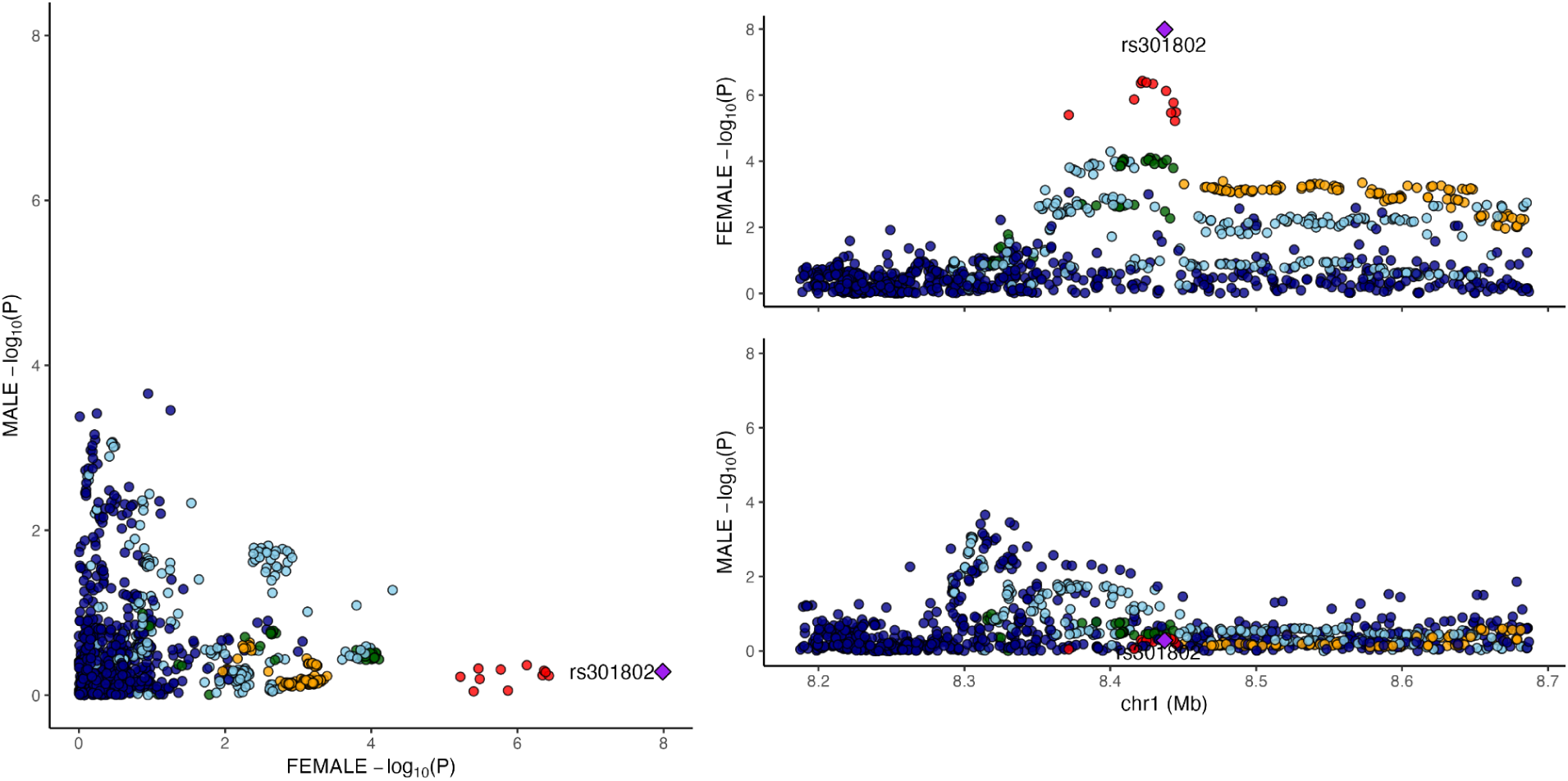
LocusCompare plot of the *RERE* locus (rs301802).

**Supplementary Figure 4:**
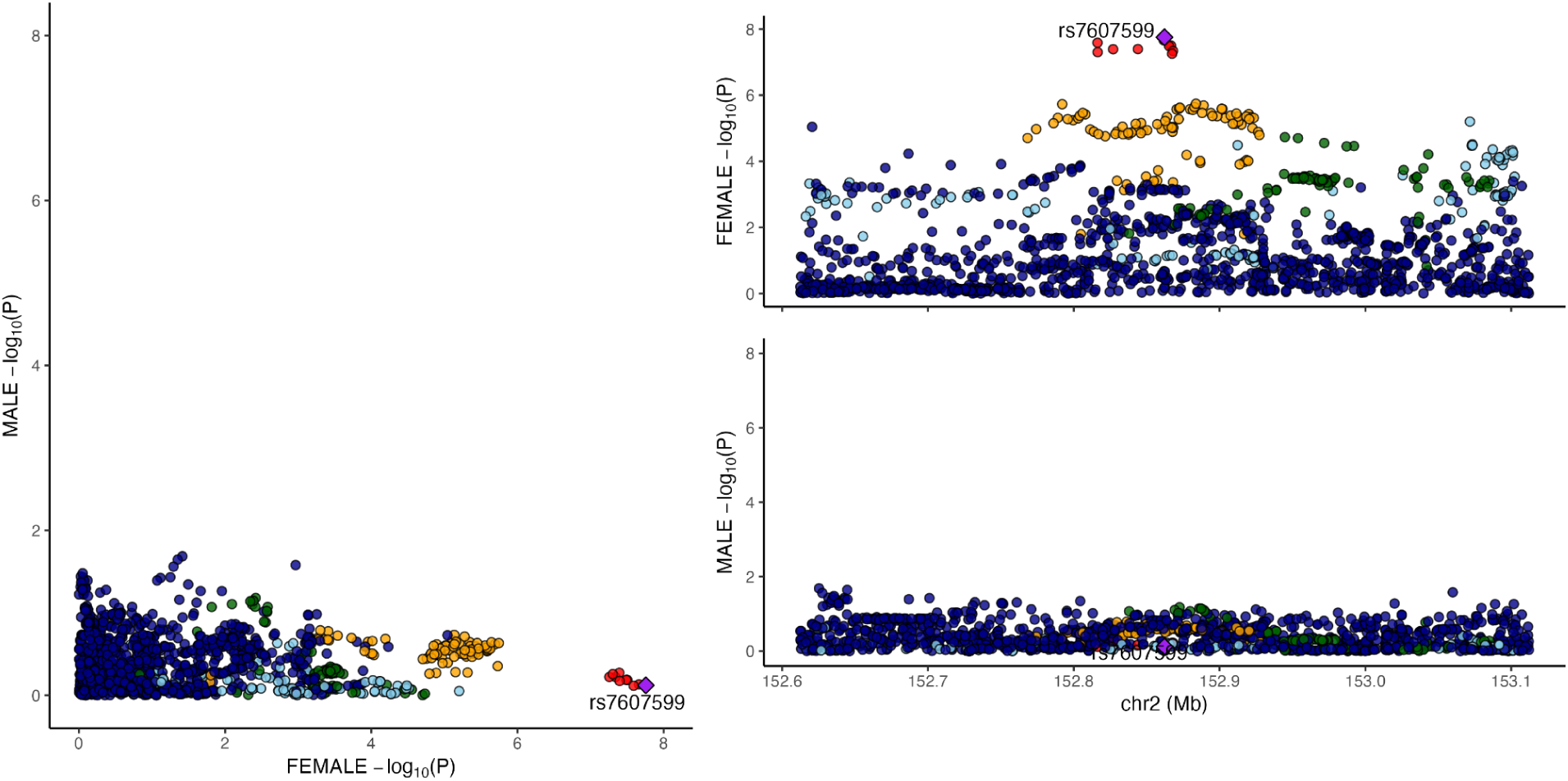
LocusCompare plot of the *ARL6IP6* locus (rs7607599).

